# Dengue Virus Serotype 2 Cosmopolitan C Genotype Reemerges with a New Subclade in Southwest Region of Bangladesh

**DOI:** 10.1101/2023.12.24.23300504

**Authors:** Md. Ali Ahasan Setu, Prosanto Kumar Das, Toukir Ahammed, Shuvo Saha, Adib Hasan, P K Shishir Kumar, Samiran Das, Tanvir Ahamed, KM Amran Hossain, Hassan M. Al-Emran, M. Anwar Hossain, Iqbal Kabir Jahid

**Affiliations:** Department of Microbiology, Jashore University of Science and Technology; Genome Center, Jashore University of Science and Technology; Department of Genetic Engineering and Biotechnology, Jashore University of Science and Technology; Department of Nutrition and Food Technology, Jashore University of Science and Technology; Department of Physiotherapy and Rehabilitation, Jashore University of Science and Technology; Department of Biomedical Engineering, Jashore University of Science and Technology

**Keywords:** Dengue serotype 2, Bangladesh, New Subclade, Cosmopolitan C, Phylogenetic tree

## Abstract

In 2023, the Dengue virus (DENV) outbreak infected over 0.3 million cases and 1500 deaths in Bangladesh. Although the the serotype and genotype data were unavailable. Our study conducted serotyping and genomic surveillance in four districts of Southwest Bangladesh between September and October 2023. The surveillance data from 2019 to 2023 extracted from the Directorate General of Health Services in Bangladesh indicated a significant increase of Dengue infections in 2023, particularly during September-November. The two-layered hypothesis examination confirmed that, despite endemic months, 2023 dengue outbreak had a higher morbidity rate compared to previous years (2019-2022) in Southwest of Bangladesh. Serotyping and E gene sequence analysis of 25 randomly selected positive samples reveals that DENV-2 was the sole serotype circulating in this region during the study period. Genomic analysis exposed a new subclade of DENV-2, classified under Cosmopolitan genotype within C clade, distinct from previous years Bangladeshi variants until 2022. This subclade, possibly migrating from India, might be emerged during COVID-19 pandemic years and exhibited higher morbidity rates, thus challenging our existing mitigation strategies. This investigation provides valuable insights for public health interventions and underscores the importance of continuous genomic surveillance in managing Dengue outbreaks.

## 1. BACKGROUND

Antigenically distinct Dengue virus serotypes (DENV-1 to 4) are further subdivided into multiple genotypes based on the sequence variations in envelope (E) gene and named according to their geographic region such as Asian I, Asian II, Asian/American, American, Cosmopolitan, and sylvatic. However, some genotypes have spread all over the world and substantially contributed to the global dengue burden and mortality. Cosmopolitan genotype of DENV-2 is the most widespread and genetically heterogeneous of all (Yenamandra et al., 2021). In Bangladesh, although sequence data is limited, DENV2 was prevalent in 2017 and 2018, whereas DENV3 was prevalent in 2019 until August 2022(Rahim et al. 2023). However, there was no report published on circulating DENV serotypes/genotypes data about ongoing 2023 outbreak in Bangladesh where more than 1500 died so far (MIS, 2023). Thus, monitoring of transmission dynamics facing a challenge of implementation of effective DENV mitigation strategies. This study endeavors to conduct genomic surveillance, providing valuable insights into the genotypes in the southwest region of Bangladesh.

## 2. METHODS AND MATERIALS

The study conducted a cross-sectional survey of dengue positive cases between September and October, 2023 in four districts of southwest Bangladesh, Jashore, Magura, Narail and Jhenadha. The genome-sequencing were randomly selected positive samples (n=5) of each week during the study period. Additionally, the study collected 30 months (2019-2023) surveillance data in those districts retrieved from the Directorate general of health services (DGHS) to perform morbidity trend analysis (https://dashboard.dghs.gov.bd/pages/heoc_dengue_v1.php).

The study was approved by the ethical review committee (ERC) of Jashore University of Science and Technology (JUST), Bangladesh (ERC no: ERC/FBST/JUST/2021-62), and all experiments were performed in the Genome Center, JUST according to the relevant guidelines and regulations. During September - October 2023, 600 suspected dengue fever patients (both hospitalized and outdoors) were selected as the study subjects. The participants were informed about the study and interviewed through a questionnaire to obtain socio-demographic and health-related data. After obtaining informed written consent from the subjects, left-over blood samples were collected from the pathology lab of different government hospital and private diagnostic centers. The blood samples were transported immediately to the Genome Center under stringent cold-chain conditions. The sera were separated through centrifugation, and a portion of the sera was stored at -40°C for further use. The other portion of the sera was used to perform Dengue NS1 antigen test and viral RNA extraction. The NS1 antigen test was performed using the Dengue NS1 Rapid Test Kit (Hangzhou Frenovo Biotech Co., Ltd., China). Viral RNA was extracted using the QIAamp Viral RNA Mini Kit (Cat. No. 52906, Qiagen, Hilden, Germany). After extraction, the DENV RNA serotyped using Molaccu Dengue Virus qPCR Detection Kit (Cat. No. 01.09.21.16.32.02, Zybio Inc., China) in qTOWER^3^G RT-PCR instrument (Analytic Jena, Germany) following manufacturer instructions. Afterward, cDNAs were prepared from the DENV2 positive RNA using SuperScript™ III First-Strand Synthesis kit (InvitrogenTM, Thermo Fisher Scientific, USA). Two overlapping fragments of the DENV2 E gene was amplified by using d2s1C-CC, DEN2 CC-1R, DEN2 CC 1F, & d2a18-CC primers (Cruz et al., 2016) and verified through electrophoresis. Sanger sequencing was performed in Applied Biosystems SeqStudio genetic analyzer using BigDye™ Terminator v3.1 as per the optimized protocol (Islam et al., 2021). All forward and reverse ab1 files from a sequencing run were simultaneously quality trimmed in Chromas and assembled in Unipro UGENE followed by genotyping in GISAID DengueServer (https://www.epicov.org/epi3/frontend#1754f5). Additional non-sylvatic DENV 2 genome sequences were retrieved from the publicly available database (NCBI Virus) and aligned with the sequenced data of this study using MAFFT.

A total of 595 sequences of DENV-2 E gene (1485bp in length) were used for phylogenetic analysis. We included 25 samples from the clinical study and 570 samples from NCBI virus database (year 2000 to 2023). We took all five genotype (Asian-I; n= 4, Asian-II; n= 2, Asian-American; n= 3, American; n= 1, Cosmopolitan; n= 585) for this analysis. Phylogenetic tree were inferred from the multiple sequence alignment using maximum likelihood method generated using IQ-TREE2. The best fit model TIN2+F+I+G4 was selected using ModelFinder and an ultrafast bootstrap with 100000 replicates was calculated (Katoh et al., 2002; Minh et al., 2020). The phylogenetic tree was visualized using i-Tol (Letunic & Bork, 2021).

The cases were analyzed through Statistical package of social sciences (SPSS) version 20. Normality test was performed by Kolmogorov–Smirnov test & Shapiro–Wilk test, and found a non-normal distribution (P<.0001). Descriptive analysis was presented by median and interquartile range (IQR). Our first hypothesis was the trend of seasonality of morbidity, we performed Kruskal–Wallis H test; second hypothesis was increased morbidity at 2023 compared to 2019-23, examined by Mann–Whitney U test (Faruk et al., 2022). The morbidity trend analysis was presented through line graph and statistically tested by Mann-Kendall trend analysis (Faruk et al., 2023). The alpha value of the study was P<.05.

## 3. RESULTS

The median morbidity cases of dengue between 2019 and 2022 were 34 (IQR 487), and in 2023 were 64 (IQR 2478). The highest median cases were observed in September (2129), October (1134) and November (512). Figure 1(A) illustrates the overall cases between 2019 and 2023 according to the distribution of months. The seasonal increase of cases (September-November) was found significant (P<.05) to be predicted as a factor. Further analysis found a significant (P<.05) mean difference in yearly cases in 2023 compared to 2019-2022. Finally, a Mann-Kendall trend analysis (Faruk et al., 2022) between monthly cases of 2019-2022 versus 2023 and found a statistically significant (P<.05) trend (Figure 1B) of increased cases in 2023 compared to 2019-2022.

**Figure 1:**
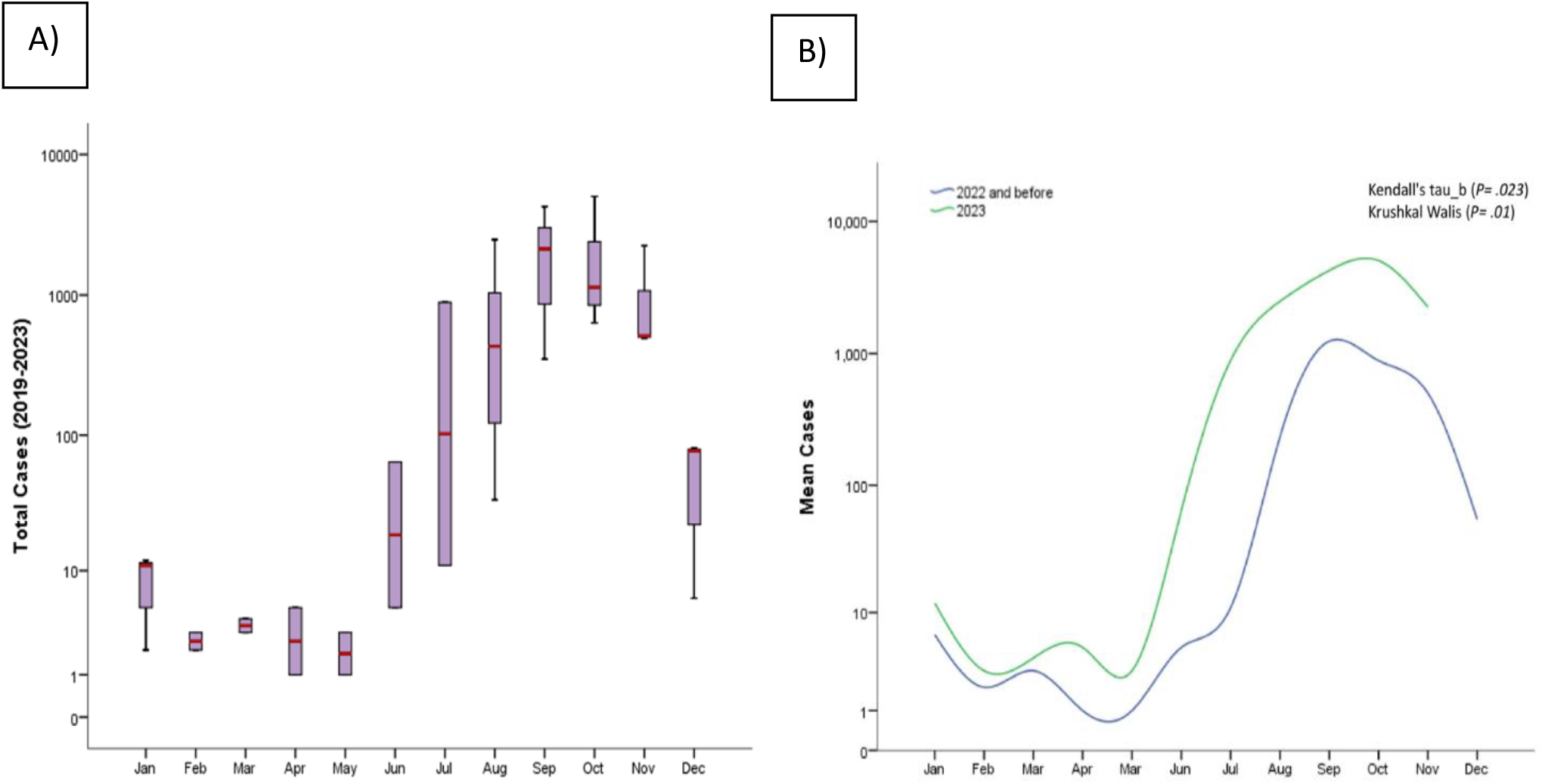
(A) Seasonal variation of Dengue cases of southern part of Bangladesh (2019-2023); (B) Month-wise Mann-Kendall trend analysis (2019-2022 versus 2023) (Data source: (https://dashboard.dghs.gov.bd/pages/heoc_dengue_v1.php)

All 595 sequences used in this study were genotyped using GISAID Dengue server GISAID. All of 25 sequences of this study clustered together in a sub-clade C of Cosmopolitan genotype as showed in Figure 2. DENV-2 Bangladeshi variants from 2006 to 2017 (n = 16) were Cosmopolitan within B clade, from 2017 to 2018 (n = 55) were Cosmopolitan C clade, whereas in 2017 the variants were in both Clade B and Clade C (Figure 2). Previous study showed that Bangladesh had no cosmopolitan sequences from 2020 to 2022 (Hossain et al., 2023). The Bangladesh 2023 subclade, we found in this study were Cosmopolitan C clade. This Bangladeshi 2023 subclade of Cosmopolitan C are genetically distant from previous Bangladeshi variants.

**Figure 2:**
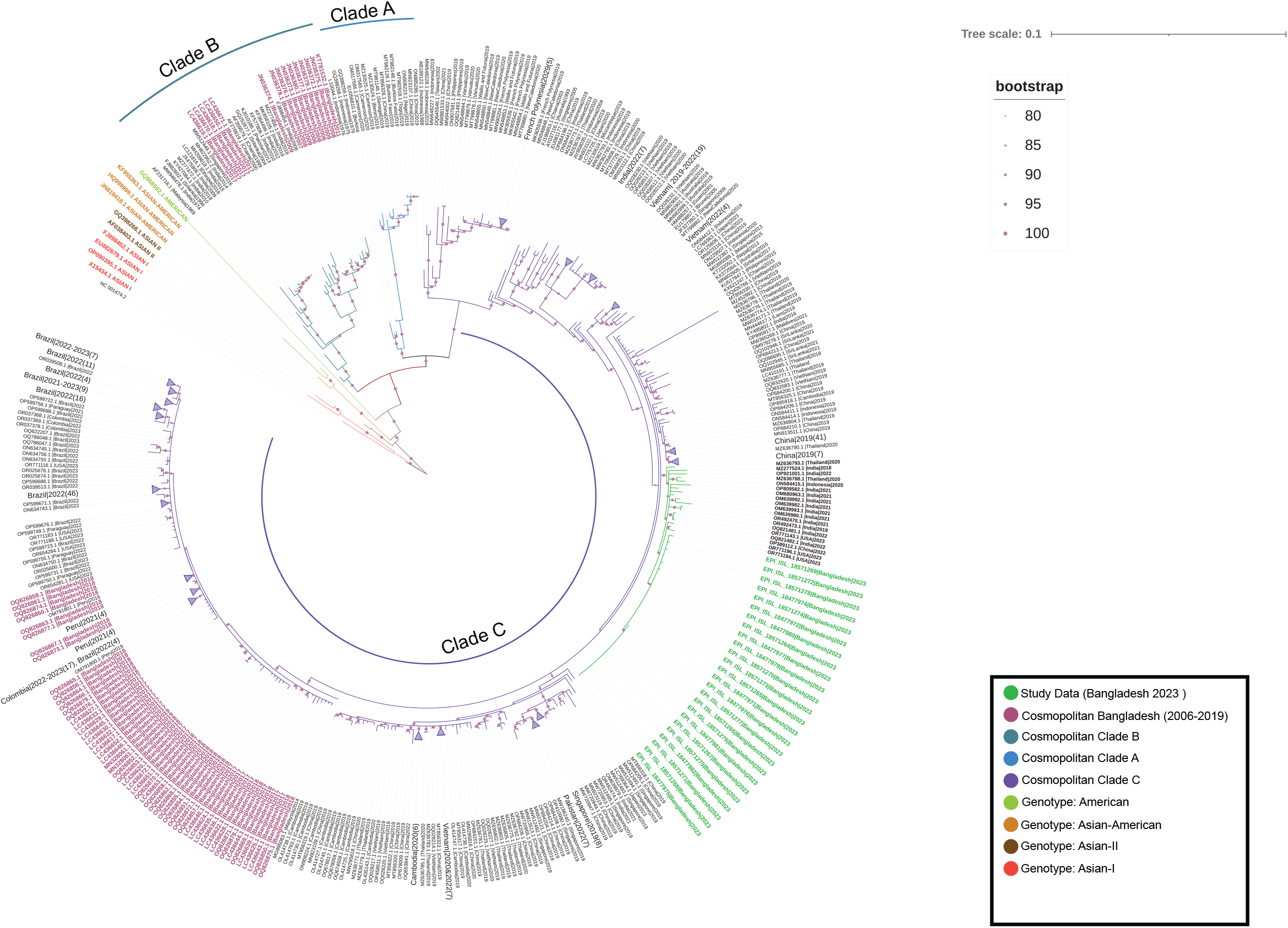
A maximum likelihood phylogenetic tree was constructed using the W-IQ-TREE program in ModelFinder with ultrafast bootstrapping (UFBoot) and 100000 replicates. TIN2+F+I+G4 was selected as the best-fit model. The dataset includes DENV-2 E-gene sequences obtained in the current study, highlighted in green for the year 2023. Additionally, DENV-2 sequences from Bangladesh spanning the years 2000 to 2022 are represented in pink. The cosmopolitan clade (A, B, and C) is demarcated by arc and corresponding colors. Each sample name comprises the accession number, country, and reported year of the sequence. Branches on the phylogenetic tree are annotated with UFBoot support values, with values exceeding 90% being highlighted.

## 4. DISCUSSION

Dengue causes a wide range of clinical manifestations, from asymptomatic or mild fever to potentially fatal diseases, such as dengue hemorrhagic fever or dengue shock syndrome. In response to dengue infection, cytokines such as IFN αβ, IL-6, IL-8, MIF, TNF α, IFN γ, Il 10, and VEGF can regulate viral replication and direct immune cells to the infection site. If this response fails, viremia develops and an amplified cytokine response leads to disease severity and affects other organs (Srikiatkhachorn et al., 2017). Secondary infection with a heterotypic serotype or genotype may elicit a prompt immune response due to the antibody-dependent enhancement (ADE) and increase disease severity (Al□Emran et al., 2023). Dengue outbreaks in Bangladesh were caused by several serotypes and genotypes in recent years including during the Covid-19 pandemic (Al-Emran et al., https://papers.ssrn.com/sol3/papers.cfm?abstract_id=4401701.; Rahim et al., 2023; Sarkar et al., 2023). The two-layered hypothesis examination proved that, despite endemic months, 2023 dengue outbreak had a higher morbidity rate compared to previous years (2019-2022) in Southwest of Bangladesh. In 2023,, more than 0.3 million cases were infected and 1500 people died in all over Bangladesh (MIS, 2023). Our study that the dengue epidemic in 2023 was caused by DENV-2 Cosmopolitan genotype that evolved as a new subclade among C clade. The outbreaks of 2017 and 2018 in Bangladesh were also caused by DENV-2 Cosmopolitan genotype within B and C clade (Hossain et al., 2023), although the 2023 subclade were not resembled to any previous Bangladeshi C clade variants (2017, 2018 and 2019), suggesting a shift of new subclade. This 2023 subclade was more similar with India (n = 12, 2018-2021), China (n =1, 2022), Indonesia (n =1, 2020), Thailand (n =2, 2020), and USA (n = 3, 2023) (Figure 2). The study area is connected with Petropole of India and Benaople the biggest land port of Bangladesh. Therefore, we assumed that this sub clade might migrate from India which had survival advantages over time. The possibility of cross-border transmission, as the isolates identified in our study closely aligned with Indian isolates identified in 2022, showing a robust bootstrap support of 99%. Our study area shares multiple borders with India, including the Benapole Land Port, the largest land port in Bangladesh.

All those variants were emerged during COVID-19 pandemic. Another study of our research group reported that the new variant might emerge from the coinfection cases with COVID-19 and causing more severity and mortality (Al-Emran et al., n.d. https://papers.ssrn.com/sol3/papers.cfm?abstract_id=4401701). The emergence of new subclade took place due to the high rate of viral replication and mutation, combined with selective pressure from the host immune responses and coinfection with other viruses. The emergence of new subclade and their potential impact on the ongoing outbreak further highlights the need for the research on coinfection and their potential effects on viral evolution and pathogenicity, and the dynamics of the cross-border spread.

## AUTHOR CONTRIBUTIONS

*Conceptualization*: M.A.A.S., I.K.J.; *Experimental Methodology*: M.A.A.S., I.K.J., P.K.D., T.A., S.S.; *Investigation*: M.A.A.S., M.A.H., I.K.J.; *Resources*: M.A.H., I.K.J.; *Sample Collection*: P.K.D., T.A., S.S., A.H., S.K.P.K., S.D., Tanvir. A.; *Experimental Analysis and Interpretation*: M.A.A.S., P.K.D., T.A., S.S. A.H., S.K.P.K.; *Validation*: I.K.J., M.A.H.; *Statistical Analysis*: K.A.H.,; *Writing, Original draft preparation*: M.A.A.S., P.K.D., I.K.J.; *Writing, Review and Editing*: I.K.J., H.A.E.; *Funding Acquisition*: I.K.J., M.A.H. All authors have read and agreed to the published version of the manuscript.

## ACKNOWLEDGMENTS

This study was supported by the Jashore University of Science and Technology grant no: *23-FoBST-06* though University Grant Commission, Bangladesh. Funding source has no role for design, collection the data, analysis and approval of manuscript.

## DATA AVAILABILITY STATEMENT

Dengue type 2 gene E that had been obtained from Sanger sequencing had been deposited in the GSAID (https://gisaid.org/). Other genetic data used in the study have been collected from NCBI (https://www.ncbi.nlm.nih.gov/). Accession number of all sequence data are available in supplementary table S1.

